# Tau-Neurodegeneration *mismatch* reveals vulnerability and resilience to comorbidities in Alzheimer’s continuum

**DOI:** 10.1101/2023.02.12.23285594

**Authors:** Xueying Lyu, Michael Tran Duong, Long Xie, Robin de Flores, Hayley Richardson, Gyujoon Hwang, L.E.M. Wisse, Michael DiCalogero, Corey T. McMillan, John L. Robinson, Sharon X. Xie, Murray Grossman, Edward B. Lee, David J. Irwin, Bradford C. Dickerson, Christos Davatzikos, Ilya M. Nasrallah, Paul A. Yushkevich, David A. Wolk, Sandhitsu R. Das, Alzheimer’s Disease Neuroimaging Initiative

## Abstract

Variability in the relationship of tau-based neurofibrillary tangles (T) and degree of neurodegeneration (N) in Alzheimer’s Disease (AD) is likely attributable to the non-specific nature of N, which is also modulated by such factors as other co-pathologies, age-related changes, and developmental differences. We studied this variability by partitioning patients within the Alzheimer’s continuum into data-driven groups based on their regional T-N dissociation, which reflects the residuals after the effect of tau pathology is “removed”. We found six groups displaying distinct spatial T-N *mismatch* and thickness patterns despite similar tau burden. Their T-N patterns resembled the neurodegeneration patterns of non-AD groups partitioned on the basis of z-scores of cortical thickness alone and were similarly associated with surrogates of non-AD factors. In an additional sample of individuals with antemortem imaging and autopsy, T-N mismatch was associated with TDP-43 co-pathology. Finally, T-N *mismatch* training was then applied to a separate cohort to determine the ability to classify individual patients within these groups. These findings suggest that T-N *mismatch* may provide a personalized approach for determining non-AD factors associated with resilience/vulnerability to Alzheimer’s disease.

## Introduction

Alzheimer’s disease (AD) is heterogenous in the age of onset, course, cognitive and behavioral phenotype, and the presence of underlying co-pathology^1–4^. In particular, concomitant pathologies are present in the vast majority of individuals with AD, such as cerebrovascular disease and/or other degenerative pathologies, including TAR DNA-binding protein 43 (TDP-43) and alpha-synuclein proteinopathies ^2,3,5,6^. Alzheimer’s also occurs in the context of the aging brain for which there is variability in age-related changes that also may influence AD clinical presentation and rate of decline. Resilience factors also appear to influence outcomes and degree of pathology associated with clinical status^7–10^. This heterogeneity, as well as a lack of well-validated markers of these non-AD influences, is a substantial challenge in the development and application of AD targeting therapeutics. As we appear to be entering a new age of disease-modifying therapies, there remains debate about who might benefit most from these interventions relative to their associated risk and to what extent specifically targeting AD-related pathology can be expected to slow decline in the context of common co-pathologies. Precision medicine approaches that categorize individuals with regard to both AD and non-AD contributions of cognitive impairment are essential as we move therapies into practice, as well as for stratification in intervention studies to advance these therapeutics.

The accumulation of amyloid plaques (Aβ) and tau neurofibrillary tangles (NFT) are the two hallmark pathological features of AD. A long history of postmortem studies^11–13^ and, more recently, work with PET imaging have supported the hypothesis that NFTs are more tightly linked to downstream neurodegeneration than amyloid plaques^14–18^. Thus, expected neurodegeneration due to AD may be largely explained by the local presence and amount of tau pathology to a first approximation. As such, measures of tau-based NFTs may provide a metric of the degree to which AD is contributing to neurodegeneration relative to other factors in any individual patient.

This idea is implicit in the National Institute on Aging and Alzheimer’s Association (NIA-AA) recently proposed AT(N) research framework^19^, which assigns individuals based on the dichotomous presence or absence of amyloid plaques (A), aggregated tau (T) and neurodegeneration (N). Given assumptions about the orderly evolution of AD from A→T→N, N in the absence of T pathology is assumed to represent non-AD pathophysiology regardless of the presence of amyloid. However, the AT(N) framework does not account for either continuous or spatially varied T-N relationships. Indeed, neurodegeneration is not specific to tau as other pathologies also contribute to neurodegeneration^5,20^. Brain resilience due to protective factors may also influence the relationship of T to N^7–10^. Thus, a measure of spatial and continuous variation of the T-N relationship may be a valuable complement to the AT(N) framework.

In the current work, we exploited the non-specific nature of neurodegeneration to account for non-AD processes by quantifying the degree and spatial pattern of deviation from the expected level of N for a given level of T, assuming a linear relationship. In essence, we are “regressing out” the effects of AD on brain structure which should result in patterns of relative atrophy associated with these non-AD factors. In other words, regional patterns of T-N discordance may reveal groups associated with specific potential co-pathologies (e.g. greater medial temporal atrophy with concomitant TDP-43) or types of resilience. To capture these patterns, we employed data-driven clustering based on regional T-N *mismatch*. We hypothesized that “vulnerable” groups with more N than expected for T would likely be associated with non-tau pathologies. Resilient groups on the other hand may be associated with protective factors, including greater brain reserve, such that they have less N than expected for T.

The current analysis builds on our prior results^21^ for development of a scalar T-N *mismatch* metric in a set of amyloid positive (A+) participants using tau PET for measuring T and gray matter thickness from MRI for measuring N (Das et al., 2021), which we also explored using ^18^F-fluorodeoxyglucose PET as the N measure^22^. We previously found meaningful associations between a T-N *mismatch* metric and a number of factors, including age, white matter hyperintensity burden and cognition. However, specific drivers of regional T-N mismatch are still yet to be determined and requires additional studies to more closely link with specific non-AD processes. The current paper takes further steps enumerated below to understand T-N mismatch and explore its underlying factors through both *in vivo* and *ex vivo* analyses.

(1) Our hypothesis is that regional T-N mismatch identifies vulnerability and resilience driven by non-AD factors that are common in AD and non-AD patients. We therefore compared the groups of T-N mismatch to amyloid negative (A-) symptomatic patients partitioned into data-driven groups on the basis of relative cortical thickness, using control referenced z-scores (N_Z_ groups). As T-N mismatch can be conceptualized as reflecting non-AD neurodegeneration after removing the effect of AD, we predicted that we would find spatially corresponding N_Z_ groups. (2) We further predicted that associations with surrogates of non-AD factors (e.g. white matter hyperintensities, advanced or slowed brain aging, TDP-43 pathology) would be associated with T-N groups and that similar patterns of associations would be found in corresponding N_Z_ groups. (3) Given comorbidities may synergistically interact with AD and contribute to cognitive impairment, we predicted that T-N groups would also differ in longitudinal cognitive decline. (4) Finally, to evaluate the potential application of this approach on individual classification in the spirit of “precision medicine”, we determined group membership of individuals in a second cohort.

## Results

### T-N groups were distinct despite similar AD severity

Based on T-N residuals in 104 gray matter regions, we grouped 184 A+ symptomatic (MCI/dementia) participants from the Alzheimer’s Disease Neuroimaging Initiative (ADNI) using hierarchical clustering^23^. Six T-N groups were determined to be optimal using the elbow method^24^ and dendrogram structure. The clinical characteristics of T-N groups are described in Table I. These groups differed in age, proportion with mild cognitive impairment (MCI) versus AD dementia diagnosis, and degree of cognitive impairment. The groups did not differ in inferior temporal (IT) mean ^18^F-flortaucipir uptake (*P=0*.*329*), a surrogate for AD severity^25,26^. Average maps of T-N linear regression residuals across brain regions of interest (ROIs) are represented visually for the six groups in Figure 1. The group with the largest number of participants (n=94) displayed low overall residuals; as such, we labeled this group “*canonical*”. Two groups with greater neurodegeneration than expected given their level of tau (N>T, negative residuals) were labeled “*vulnerable*.” One of these groups had N>T mostly in temporal/limbic regions, which we labeled as “limbic vulnerable” (n=25); the other group (n=18) displayed widespread N>T throughout the cortex encompassing both limbic and neocortical regions and was labeled “diffuse vulnerable”. In addition, there were two T-N groups with less neurodegeneration than expected given the level of tau (N<T, positive residuals) and we labeled them as “*resilient*.” One (n=16) displayed N<T in the temporal regions with extension to temporal-occipital cortex was denoted as posterior-temporal occipital (PTO) resilient. We labeled the other group (n=26) showing distinct N<T in lateral and medial temporal cortex and parts of prefrontal cortex as anterior-temporal (AT) resilient. A final small group (n=5) with N<T especially along the motor cortex, but N>T in temporal/limbic region, was labeled “*mixed*.”

**Table I.** Characteristics of distinct groups via T-N *mismatch* clustering for 184 A+ symptomatic patients. Overall group effects are tested using the Kruskal-Wallis test for categorical variables (sex, MCI/Dementia) and linear regression for continuous variables (age, years of education, MMSE, CDRSB and inferior temporal (IT) tau SUVR). The baseline cognitive scores (Mini-Mental State Exam (MMSE)^27^, Clinical Dementia Rating Sum of boxes (CDRSB)^28^) and IT Tau SUVR was compared with age, sex and years of education as covariates. The mean (SD) is shown for age, years of education, MMSE, CDRSB and IT tau SUVR. Pairwise comparisons of these variables between T-N groups were obtained. Only significant pairwise comparisons between T-N groups the Group1 (canonical) were marked in the table. P-values were adjusted by Bonferroni multiple comparison correction (*\*P<0*.*05, **P<0*.*01, ***P<0*.*001)*.

| Group (n) | Description | Age | Sex (F/M) | Diagnosis (MCI/Dem) | Educ (SD) | MMSE (SD) | CDRSB (SD) | IT Tau SUVR (SD) |
| --- | --- | --- | --- | --- | --- | --- | --- | --- |
| Group1 (94) | Canonical | 76.4 (7.3) | 44/50 | 59/35 | 15.6 (2.5) | 25.8 (3.3) | 2.97 (2.1) | 1.48 (0.41) |
| Group2 (25) | Limbic Vulnerable | 80.2 (6.8) | 11/14 | 8/17 | 16.6 (2.5) | 23.1** (3.9) | 6.21*** (4.4) | 1.48 (0.35) |
| Group3 (18) | Diffuse Vulnerable | 79.4 (9.2) | 3/15 | 5/13 | 15.5 (3.1) | 23.4* (5.2) | 4.62 (3.2) | 1.42 (0.44) |
| Group4 (16) | Posterior-Temporal Occipital Resilient | 70.1* (7.9) | 12/4 | 12/4 | 15.9 (2.7) | 26.4 (3.9) | 3.50 (3.5) | 1.54 (0.43) |
| Group5<br>(26) | Anterior-Temporal<br>Resilient | 73.1<br>(7.8) | 9/17 | 22/4 | 16.4<br>(2.7) | 26.4<br>(3.8) | 1.70<br>(1.5) | 1.39<br>(0.3) |
| Group6<br>(5) | Mixed | 73.7<br>(6.6) | 2/3 | 2/3 | 15.0<br>(3.0) | 25.0<br>(4.6) | 2.10<br>(1.7) | 1.66<br>(0.39) |
| Group<br>Diff. | -- | $P<0.001$ | $P=0.024$ | $P<0.001$ | $P=0.435$ | $P<0.001$ | $P<0.001$ | $P=0.329$ |

**Figure 1.**
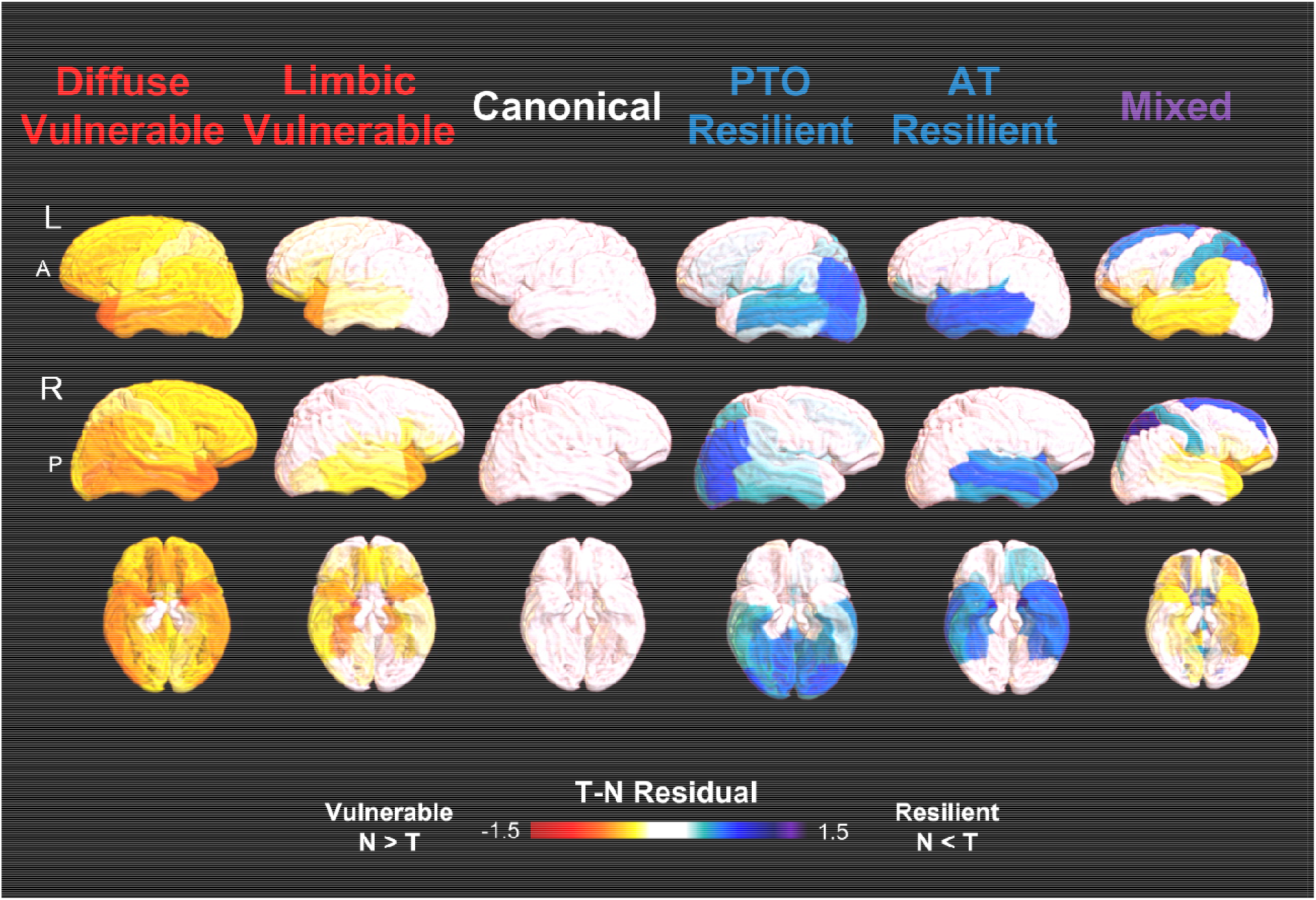
Average ROI-wise residual maps representing spatial T-N relationships for identified T-N groups among A+ symptomatic patients from ADNI via T-N *mismatch*: canonical (close to 0 residuals, N∼T), vulnerable (negative residuals, N>T), resilient (positive residuals, N<T).

To determine in a more granular manner whether the T-N groups differed in tau burden, we assessed regional tau SUVR in representative limbic and cortical regions. Figure S1 further demonstrates that the groups did not differ in ^18^F-flortaucipir uptake across limbic/cortical regions (*P’s>0*.*05*). However, the between-group thickness covaried with regional tau, age, and sex differed in the same representative regions, supporting the contention of their “mismatch” status (Figure S1).

### Clustering A-symptomatic patients based on patterns of neurodegeneration alone reveals groups similar to those from T-N mismatch

We also performed data-driven clustering of 159 MCI/dementia ADNI participants who were A-using regional z-scores of cortical thickness relative to A-cognitively normal adults, referred to as “N_Z_ clustering”. The same ROIs were used as for the analogous T-N mismatch-based clustering. We determined that five N_Z_ groups were optimal in this case. Notably, the N_Z_ group patterns of average z-score thickness resembled the residual patterns of the vulnerable T-N groups (Figure 2A). In particular, we found a group (n=10) that had more negative z-scores in limbic regions, a pattern that resembled the T-N limbic vulnerable group, and we therefore labeled it as “limbic atrophy”. Two of the groups displayed relatively diffuse low z-scores that corresponded to the T-N diffuse vulnerable group and were combined (“diffuse atrophy”, n=27; see Figure 2A) for additional analyses given their similar patterns (see Figure S2 for display of thickness map for these two groups separately). The largest group (n=98) displayed minimal evidence of atrophy based on the control-referenced z-scores, which we labeled it as “no-atrophy”. Finally, a group (n=24) with more positive z-scores of cortical thickness in posterior temporal and occipital regions had a very similar pattern to the residual map of the T-N PTO resilient group, and was labeled as “posterior-temporal occipital (PTO) increased thickness”.

**Figure 2.**
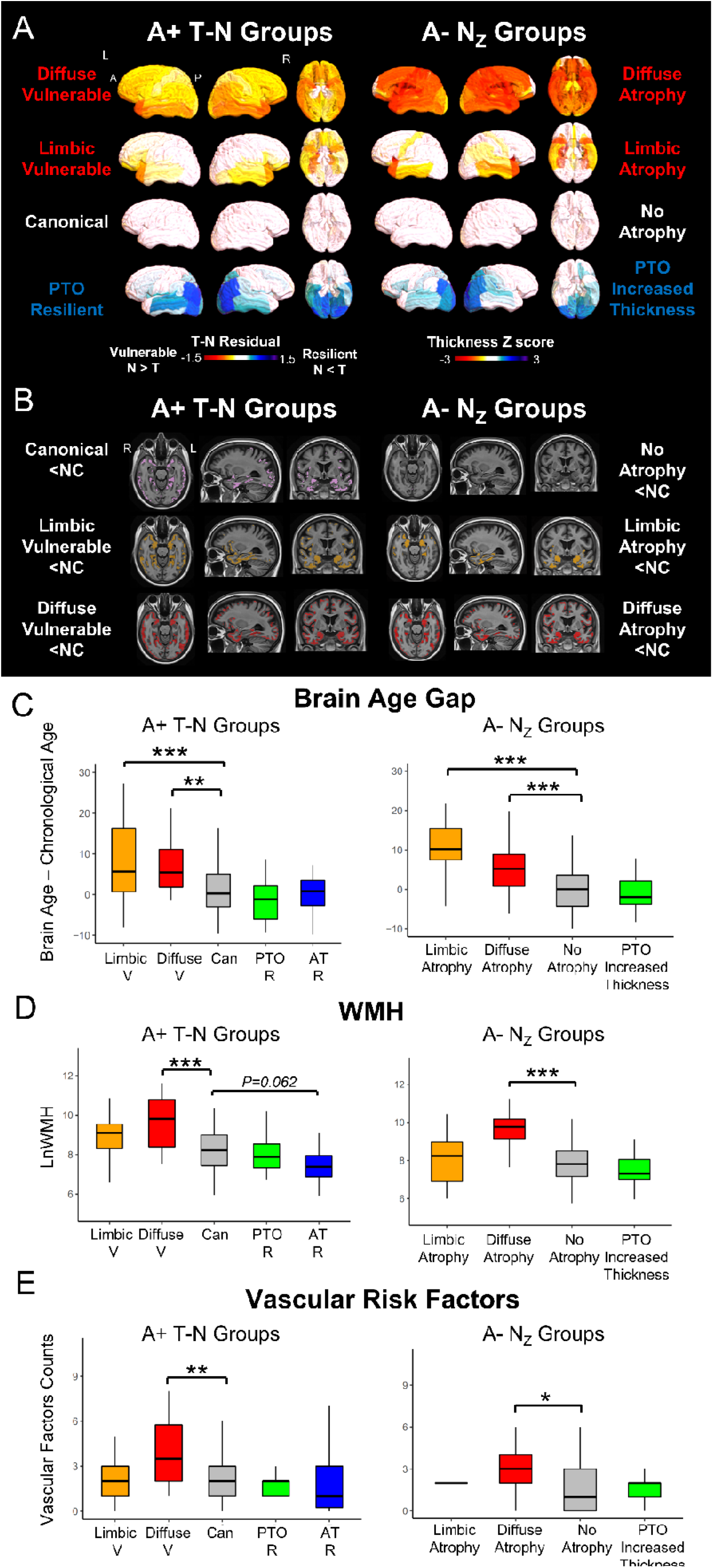
T-N groups resembled N_Z_ groups and were associated with non-AD modulators in ADNI cohort. (A) The T-N residual patterns for T-N groups were similar to regional thickness z-score patterns for identified groups among A-patients from ADNI by clustering on standardized thickness using 137 normal individuals: No atrophy (close to 0 z-score), atrophy (negative z-score), increased thickness (positive z-score). (B) Voxel-wise analyses reveal significantly less thickness of T-N groups (left) and N_Z_ groups compared with cognitively unimpaired control (NC, N=137) controlling for age and sex (*P*_*FWER*_ *< 0*.*05*). The colored areas represent significant difference with the cognitively unimpaired control for each group. (C) The between-group brain age gap (brain age – chronological age) pair-wise comparison for T-N groups (left) and N_Z_ groups (right) after covarying by age. (D) The between-group white matter hyperintensities volume comparison with age as covariates among T-N groups (left) and N_Z_ (right) groups. (E) The pair-wise comparisons of vascular risk factors for T-N groups (left) and N_Z_ groups (right). The count of vascular risk factors is the sum of participant risk factors, assayed categorically, for hypertension, hyperlipidemia, type II diabetes, arrhythmia, cerebrovascular disease, endovascular management of head/neck vessels, coronary artery disease, coronary interventions, heart failure, structural heart defects/repair, peripheral artery disease, and smoking. The mixed group was excluded in the analyses of T-N groups comparison due to its small size. For C-E, pairwise comparisons between groups were performed. Only significant comparisons with the typical groups (canonical for T-N groups, no atrophy group for N_Z_ groups) respectively were marked in the figures. Significant levels corrected by Bonferroni multiple comparison are denoted as *\*P<0*.*05, **P<0*.*01, ***P<0*.*001*.

Overall, the general overlap of spatial patterns between the residuals in the T-N mismatch groups in A+ and the cortical thickness in the N_Z_ groups in A-(Figure 2A) is consistent with the notion that T-N residuals reflect non-AD related phenomenology which may be present irrespective of amyloid status. It is not surprising that the N_Z_ group had less representation of “resilient” groups, as in symptomatic individuals without AD, there is no defined dominant neuropathology against which resilience can be measured. Aligning with the T-N groups, N_Z_ groups also varied in clinical characteristics in a similar fashion. For example, the PTO increased thickness group was younger than the no-atrophy group, and the limbic atrophy group showed poorer baseline cognition (Table S1). The N_Z_ groups did not differ in inferior temporal mean ^18^F-flortaucipir uptake values, which were generally at a level below a typical threshold for “positive” T^26^.

Next, we performed voxel-wise comparison of cortical thickness of each group from both the T-N and N_Z_ clustering results with 137 A-cognitively normal participants from ADNI. We predicted that patterns of cortical thinning would reflect, to a large extent, regional aspects of T-N mismatch patterns overlaid on typical AD effects in the T-N groups. Figure 2B shows voxels with significant differences in thickness (*P*_*FWER*_ *< 0*.*05*), controlling for age and sex, from cognitively normal individuals for both T-N and N_Z_ groups. As expected, the T-N canonical group displayed atrophy in the medial temporal lobe and posterior neocortical regions, corresponding to a “typical” pattern of AD pathology. As expected given how it was derived, the N_Z_ no-atrophy group did not differ from normal controls in cortical thickness. Alternatively, cortical thinning relative to controls was strikingly similar between T-N vulnerable and N_Z_ atrophy groups, although somewhat more extensive in the former likely stemming from the concomitant presence of AD-related neurodegeneration. The T-N limbic vulnerable and N_Z_ limbic atrophy groups both displayed prominent atrophy in temporal/limbic regions, while the T-N diffuse vulnerable and N_Z_ diffuse atrophy groups displayed more widespread atrophy throughout the temporal, parietal/occipital and frontal lobes. The T-N mixed group was excluded for voxel-wise comparison due to its small size. The T-N resilient groups and N_Z_ PTO increased thickness group did not display thickness differences with cognitively normal controls.

We also directly compared cortical thickness between groups with the respective canonical or no-atrophy groups in both T-N and A-N_Z_ groups respectively (Figure S3). T-N vulnerable and N_Z_ atrophy groups displayed significant reduction in cortical thickness relative to the respective T-N canonical or N_Z_ no-atrophy group whereas the T-N resilient and N_Z_ PTO increased thickness groups displayed regions of increased cortical thickness. No significant voxel-wise differences were observed for any of these groups in the opposite direction (vulnerable/atrophy > canonical/no atrophy, resilient/increased thickness < canonical/no-atrophy).

### T-N groups were modulated by specific non-AD factors

We sought to explore factors that may influence the degree of neurodegeneration beyond AD and, thus, may drive patterns seen in the resilient versus vulnerable groups. We compared these possible modulators in both T-N groups and N_Z_ groups, reasoning that if these groups represent the presence of concomitant non-AD pathologies, they should show a similar association with measures suggestive of these pathologies (e.g white matter hyperintensities associated with cerebrovascular disease).

In our prior work^21^, we demonstrated that relative mismatch of T and N was linked to age, as age is associated with atrophy in the absence of AD pathology. However, brain aging varies across individuals, and there are some that exhibit accelerated brain age changes for their chronological age and those that have younger appearing brains^29,30^. We reasoned that accelerated or deaccelerated brain age may be a source of vulnerability or resilience beyond chronological age and would have a similar influence of T-N and N_Z_ groups.

To assess this, we used a recently published machine learning based algorithm^31^ that uses MRI scans to infer a measure of brain age that is specifically formulated to be orthogonal to AD-related brain changes. We calculated “brain age gap,” which is the difference between the MRI-based brain age prediction and chronological age. Figure 2C plots the “brain age gap”, defined as the difference between the MRI-based brain age prediction and chronological age, for the T-N and N_Z_ groups. In the T-N groups, both vulnerable subgroups were associated with significantly greater brain age gap (brain age > chronological age) than the canonical subgroup (*p<0*.*001* for limbic vulnerable, *p<0*.*01* for diffuse vulnerable), while the PTO resilient group tended to have lower brain age gap than the canonical group, although the effect did not survive multiple comparisons correction. Brain age gap in both N_Z_ atrophy groups similarly demonstrated a significantly greater brain age gap compared to the non-atrophy group (*p<0*.*001*). The similarity of these patterns between the T-N and N_Z_ groups supports the idea that accelerated or decelerated brain aging is a factor that drives T-N mismatch which could also be present in non-AD symptomatic cases.

Another potential modulator of N outside of T is the presence of cerebrovascular disease. To determine whether our T-N groups were associated with more or less vascular disease relative to the T-N canonical group, we assessed the volume of white matter hyperintensities (WMH), a surrogate for cerebrovascular disease^32^ with age included as a covariate (Figure 2D), as well as number of vascular risk factors^32^ (Figure 2E). We found that the T-N diffuse vulnerable group had significantly higher WMH volume compared to the T-N canonical group *(P<0*.*001*), and was also associated with greater number of vascular risk factors *(P<0*.*01)* than the canonical group (Figure 2D, E). Conversely, the T-N AT resilient group tended to have lower WMH volume compared to the T-N canonical group (*P=0*.*062*). Likewise, among N_Z_ groups, the diffuse atrophy group displayed significantly higher WMH (*p<0*.*001*) and greater number of vascular risk factors (*p<0*.*05*) compared to the no-atrophy group.

### Postmortem T-N groups were associated with co-pathologies

We next attempted to determine a potential link between histological evidence of concomitant proteinopathies, TDP-43 and alpha-synuclein, with T-N mismatch groups. In particular, we hypothesized that the T-N limbic vulnerable group would display evidence of concomitant TDP-43 pathology, such as that observed in limbic-predominant age-related TDP-43 encephalopathy (LATE)^33^, given the link between LATE and greater MTL involvement in AD^34^. We used data from the Brain Bank of the University of Pennsylvania Center for Neurodegenerative Disease Research (CNDR) and antemortem MRI. Cortical thickness from antemortem MRI was again used as a measure of N, and regional semi-quantitative tau pathology ratings from postmortem histology served as a measure of T. We included 112 A+ autopsies defined by a Consortium to Establish a Registry for Alzheimer’s Disease (CERAD) score of >= 2 given that this reflects a typical threshold for detection of amyloid positivity with PET^35^. Using six cortical regions in which we had both tau pathology ratings and corresponding MRI ROIs, we defined T-N *mismatch* in each of these regions. We obtained five groups through the same clustering approach as for the *in vivo* data and compared thickness across the entire brain to the canonical group (Figure 3A). We observed similar patterns of thickness differences to what were described in the preceding *in vivo* analyses (Figure S3). We therefore denoted these groups as limbic vulnerable, diffuse vulnerable, PTO resilient and diffuse resilient, in addition to a canonical group. Demographics are presented in Table S2.

**Figure 3.**
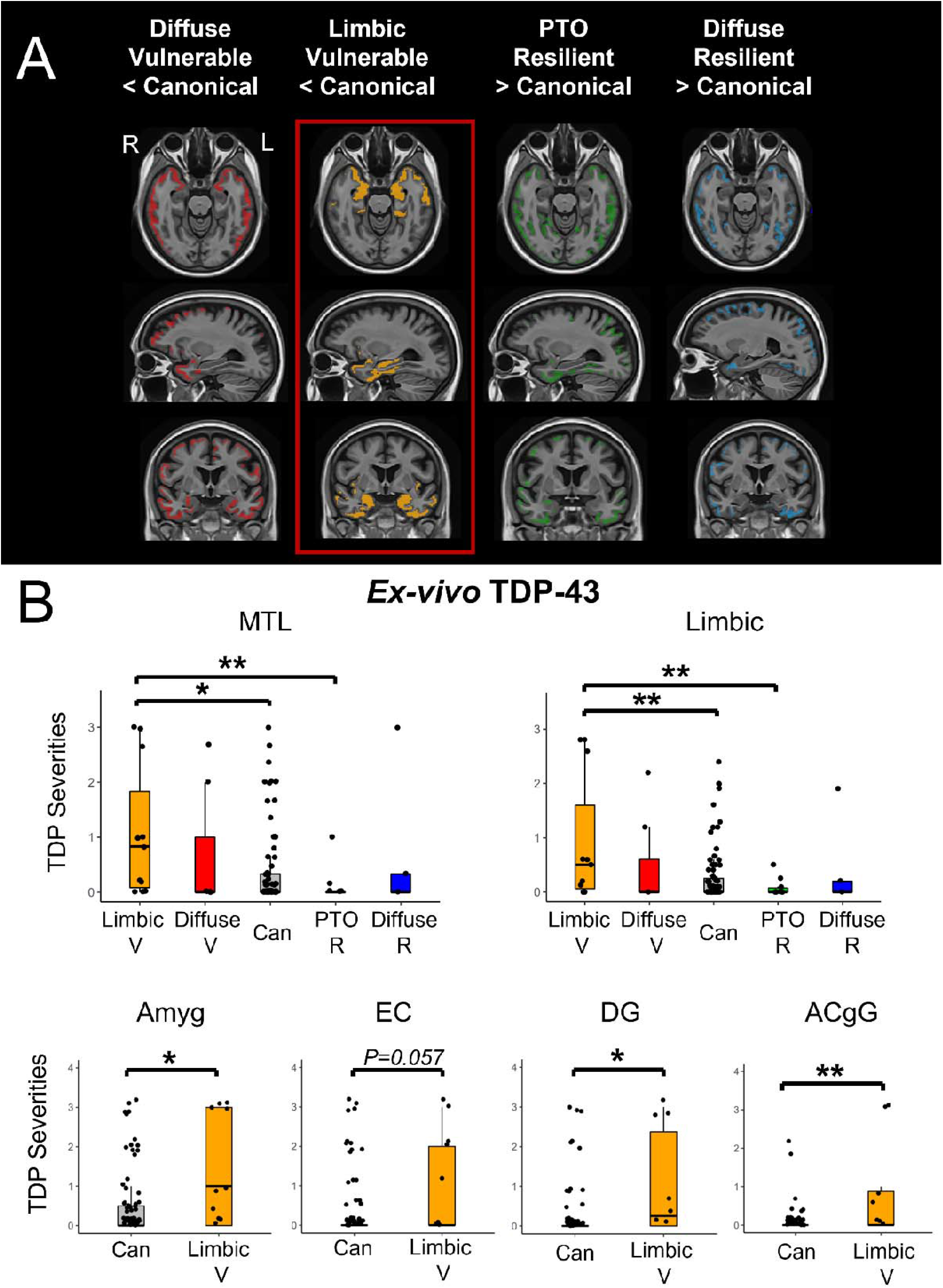
Postmortem assessment of TDP-43 on T-N groups of CNDR autopsies (A) Voxel-wise significant differences of antemortem thickness between vulnerable/resilient groups and the canonical group with *P*_*FWER*_ *< 0*.*05* for CNDR autopsies. The colored areas represent significant difference with the canonical for each group. (B) The between-group comparison *of ex-vivo* TDP-43 burden of CNDR cohort in medial temporal lobe, la limbic composite ROI as well as semi-quantitative TDP-43 severities across representative regions of early TDP-43 deposition including amygdala (Amyg), entorhinal cortex (EC), dentate gyrus (DG) and anterior cingulate gyrus (ACgG). The comparisons were not corrected by multiple comparisons given that we specifically predicted that the limbic vulnerable group would be associated with TDP-43 pathology in this case rather than performing exploratory analysis. Significant levels are denoted as *\*P<0*.*05, **P<0*.*01, ***P<0*.*001*.

We analyzed TDP-43 pathology in regions associated with early stages of LATE neuropathic change (LATE-NC) across the different T-N mismatch groups (Figure 3B). Consistent with our hypothesis, we found the limbic vulnerable group was the only group with significantly higher TDP-43 severity than the canonical group in the medial temporal lobe region (MTL, *P=0*.*015*) as well as in a composite limbic region (*P<0*.*01*). Moreover, it was associated with higher TDP-43 level in regions of early TDP-43 deposition including amygdala (Amyg, *P=0*.*01*), entorhinal cortex (EC, *P=0*.*057*), dentate gyrus (DG, *P=0*.*023*) and anterior cingulate gyrus (ACgG, *P<0*.*01*) compared to the canonical group (Figure 3B). Note that these differences did not survive correction for multiple comparisons, but were consistent with a strong a priori hypothesis. Interestingly, the PTO resilient group displayed the least amount of TDP-43 in MTL and limbic regions which was significantly lower than limbic vulnerable group (*P<0*.*01*), but its difference in TDP-43 was not significant in comparison to the canonical group (*P>0*.*05*). We also examined alpha-synuclein levels between groups specifically at amygdala, MTL and limbic composite region given early involvement in those structures. We did not find groups differences (*P=0*.*86* for Amyg, *P=0*.*79* for MTL, *P=0*.*38* for limbic composite ROI).

### T-N mismatch predicts longitudinal cognitive changes

In light of the potential role for the presence or absence of co-pathologies on longitudinal trajectory of decline, we examined differences in CDR-SB change, a measure that incorporates cognitive and functional data, among the different T-N mismatch groups from ADNI. We predicted that T-N vulnerable groups would display faster cognitive decline over time due to the possibility of comorbid pathologies while resilient groups would be expected to have slower decline. Figure 4 Left displays longitudinal CDR-SB performance for different T-N groups. The T-N vulnerable groups displayed steeper CDR-SB increases than canonical (*P<0*.*001* for limbic vulnerable group, *P<0*.*05* for diffuse vulnerable group). Alternatively, the AT resilient group tended to progress slower although not significantly so relative to the canonical group after multiple comparisons correction.

**Figure 4.**
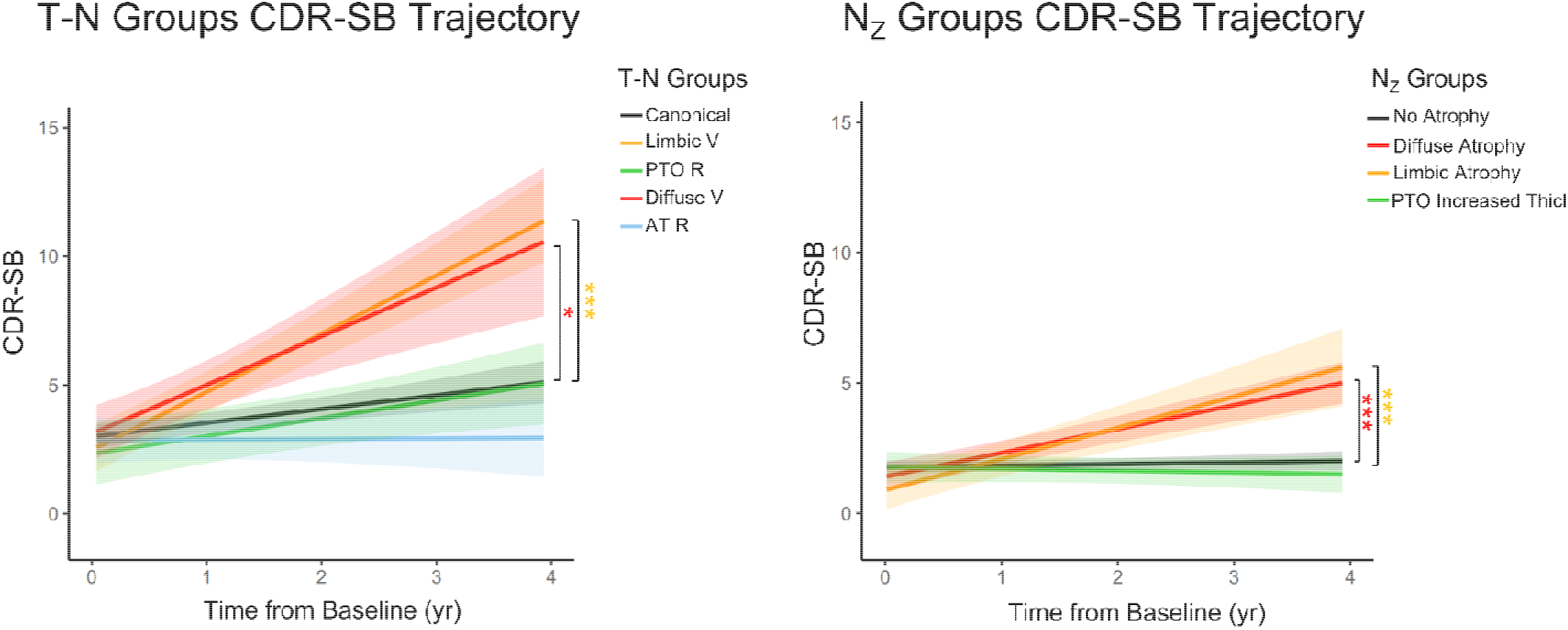
Longitudinal CDR-SB changes for all T-N groups (left) and N_Z_ groups (right) of ADNI cohort covaried with age, sex, years of education and baseline CDR-SB score. The shadow represents 95% confidence intervals. The mixed group was excluded in the analysis of T-N group comparison due to small sample size. Only significant comparisons with the typical group (canonical for T-N groups, no atrophy group for N_Z_ groups) were marked. Significant levels corrected by Bonferroni multiple comparison are denoted as *\*P<0*.*05, **P<0*.*01, ***P<0*.*001*.

We also examined longitudinal cognitive changes in the N_Z_ groups (Figure 4 Right). We predicted similar relative decline between analogous groups to the T-N analysis, but slower rates of decline due to absence of concomitant AD. Indeed, the N_Z_ atrophy groups displayed a faster rate of decline than the no atrophy group (*P<0*.*001* for limbic atrophy, *P<0*.*001* for diffuse atrophy), consistent with the expectation that non-AD pathologies would be driving decline. Moreover, the average annualized rate of change for T-N limbic vulnerable (2.48 points/year) and T-N diffuse vulnerable (2.43 points/year) groups were both greater than the rates for N_Z_ limbic atrophy (1.17 points/year) and diffuse atrophy (0.92 points/year) groups.

### Application of T-N mismatch to individual patients

To assess the transferability of T-N mismatch and its potential clinical utility on classification of individuals, we applied the T-N mismatch framework to an independent testing cohort, referred to as “AVID”, using existing T-N residual models and then inferred their T-N group identity. The testing cohort contained 71 symptomatic A+ patients, including 29 with dementia and 42 with MCI. The regional T-N residuals of each patient was obtained by projecting their T-N relationships on the existing regional T-N regressions models in the ADNI cohort. Group assignment for each individual patient was then identified by finding the ADNI T-N group with the *lowest distance* based on these imputed residuals (see Methods for details). We then grouped the patients based on inferred T-N group identity and visualized their average T-N residual maps (Figure S4). All six groups were recapitulated on the AVID dataset (Figure S4). Likewise, they were associated with clinical differences similar to that of ADNI (Table S3), including the limbic vulnerable group displaying the poorest MMSE and Alzheimer’s disease assessment scale cognitive subscale-11 (ADAS-Cog11) scores among all groups and the resilient groups generally performing better than the other groups. Despite the regression model coming from the ADNI dataset, the groups also did not differ in IT Tau SUVR, as with the ADNI analysis.

## Discussion

In the current study, we defined cognitively impaired individuals in the AD continuum based on the relationship of T pathology to N to explore the implications of variability particularly with respect to the potential role of non-AD pathologies (e.g. TDP43, vascular disease) and accelerated aging on these differences. We identified six *in vivo* data-driven groups associated with different spatial patterns of T-N relationships. The largest group was defined as “canonical”, in which the relationship between T and N was largely close to the regression line in all regions (i.e., N∼T). A limbic vulnerable group displayed greater neurodegeneration than tau (N>T) in limbic regions while a diffuse vulnerable group displayed N>T throughout the cortex. On the other hand, there were groups with relatively less neurodegeneration given tau (N<T) that we classified as posterior-temporal occipital resilient or anterior-temporal resilient based on the spatial pattern of these relationships. Finally, we also obtained a mixed group presenting both vulnerable and resilient features. Since the mean tau burden, as measured by tau PET, was similar across these phenotypic groups, these T-N residual groups do not appear to be simply reflective of AD severity per se, but we propose are driven by non-AD factors. Notably, many of the residual patterns of these T-N groups also appeared in A-symptomatic individuals partitioned based on regional gray matter thickness. As these individuals would be expected to have non-AD drivers of their symptomatic state and patterns of atrophy, similarities between these A-N_Z_ groups and the T-N groups were expected and support the hypothesis that the T-N residuals reflect the non-AD factors that contribute to neurodegeneration concomitant with AD pathology.

The vulnerability and resilience we observed in the T-N groups may be attributable to a variety of potential drivers. One potential contributor is “normal” age-related brain changes. Age itself is associated with structural changes to regional thickness and volume of grey matter which is at least partially dissociable from structural changes associated with typical AD^31,36^. As such, we previously predicted and found that age was associated with a more global measure of T-N mismatch^21^. In the current analysis, vulnerable groups do tend to be older than resilient ones, with the canonical group around the mean age of the overall cohort. However, brain age also varies across individuals, with some exhibiting more accelerated brain age for their chronological age and vice versa. Indeed, these differences may be one potential source of brain resilience and vulnerability in the context of AD and other neurodegenerative conditions. We examined this brain age gap in the T-N groups using a measure of brain age which was specifically designed to dissociate from brain changes associated with AD^31^. Using this approach, we found that both vulnerable groups have older brain age relative to chronological age compared to the canonical group. Additionally, the resilient groups tended to have relatively younger brain age than their chronological age. While this result supports the potential role of brain age beyond chronological age as a driver of vulnerability and resilience, it is worth noting that the brain age measure was not independent from non-AD pathologies which are also likely to contribute to these different groups.

Indeed, non-AD co-pathologies are very common in individuals with AD pathology and are likely important contributors to the heterogeneity of AD^1–3,5,6^. One of the vulnerable groups was referred to as “limbic” because of the greater temporal lobe, including temporal pole, and orbital frontal cortex neurodegeneration relative to tau pathology. This group also displayed generally greater cortical thinning in these regions when directly compared to the canonical group, controlling for age and sex, despite similar inferior temporal tau burden. While there are a number of non-AD related pathologies that may overlap with this region (e.g. argyrophilic grain disease), a particularly important pathology associated with limbic involvement is TDP-43, especially in the form of LATE, which is a common co-pathology with AD. LATE has previously been shown to accelerate cognitive progression and hippocampal atrophy when co-occurring with AD relative to AD alone^6,37^ although can also occur independently^33^. The pattern observed in the limbic vulnerable group is consistent with expected regional distribution of pathology and atrophy observed in TDP-43 proteinopathy ^20,33,38^. This group demonstrated a more rapid rate of cognitive decline relative to the canonical group, also consistent with prior work^33,37,39^ studying this co-pathology in the setting of AD. Moreover, the limbic vulnerable group was recapitulated in the postmortem analysis demonstrating an atrophy pattern remarkably similar to that in the *in vivo* group. Most importantly, this T-N group demonstrated higher levels of TDP-43 deposition in medial temporal lobe and limbic regions compared to other T-N groups, including the canonical group, supporting the hypothesis that TDP-43 proteinopathy drives limbic T-N mismatch.

We also found that a T-N group that we labeled posterior-temporal resilient tended to contain the least amount of TDP-43 in medial temporal lobe based on the postmortem analysis. It is possible that this group reflects individuals particularly resistant to TDP-43 pathology. Aligning with this idea, it has been argued that resilience to AD may partly depend on resistance to TDP-43, or other pathologies^40^. Given that even the canonical group in the post-mortem analysis had some degree of TDP-43 pathology and that comorbidity is much more common in AD than its absence (Figure 3B), perhaps the canonical group is actually a mixed group, and that it is those groups that were named “resilient” that don’t have co-pathology. The canonical group, rather than being “pure” AD, may contain a medium level of co-pathologies; the PTO resilient and limbic vulnerable groups then may be associated with the least and the most TDP-43 severities, respectively.

The diffuse vulnerable group was found to be associated with greater WMH volume and number of vascular risk factors^41^ compared to the canonical group when controlling for age, suggesting that cerebrovascular pathology may be a contributor to the apparent “vulnerability” in this group. Alternatively, the AT resilient group had significantly lower WMH volumes than the canonical group. While we would expect cerebrovascular pathology to have a more diffuse, or perhaps frontal, effect on cortical thinning, it is unclear why the AT resilient group largely displayed less thinning in temporal regions. There is at least some data^3,42^ suggesting that TDP-43 is more common in the setting of cerebrovascular factors such that its absence may allow for AT resilience to TDP-43. Further, aligning with TDP-43 findings above, the canonical group had evidence of an intermediate degree of vascular disease between AT resilient and diffuse vulnerable. The AT resilient, canonical and diffuse vulnerable groups may be on a continuum of cerebrovascular pathology (AT resilient < canonical < diffuse vulnerable). This again suggests that the degree of co-pathology may modulate relative resilience or vulnerability, but that “typical” AD is marked by modest degree of other pathologies, consistent with autopsy data.

Clustering based on A-thickness also produced a more diffuse atrophy group with high levels of WMHs and vascular risk factors while those with less atrophy had less evidence of small vessel disease. The fact that similar patterns were again observed in the Nz groups again supports the hypothesis that T-N mismatch is teasing out similar non-AD modulators of resilience and vulnerability.

Importantly, we found that T-N *mismatch* classification had implications for not only cross-sectional measures of cognition, but also longitudinal outcomes consistent with our prior work in a smaller cohort A+ individuals^21^. Vulnerable groups declined significantly faster than the canonical groups, likely attributable to the presence of the non-AD pathologies suggested above. In the setting of AD, tau and other pathologies may synergistically interact to accelerate cognitive impairment^6,37,39^. These findings are consistent with other work demonstrating that comorbid pathologies contribute to dementia phenotype and course^6,43,44^. Moreover, the AT resilient group demonstrated relatively little evidence of progression although not statistically different from the canonical group which had a modest rate of decline. Thus, T-N mismatch appears to have important implications for prognosis and potential stratification in intervention studies. Interestingly, the parallel N_Z_ groups displayed similar, but generally less steep decline as measured by the CDR-SB. This result is consistent with the expectation that AD plus other comorbidities result in faster decline than isolated non-AD drivers of decline^6,37,39,43^.

As further support for the robustness and potential clinical utility of this approach, we were able to make portable and straightforward inference of T-N group for individual patients from a second cohort based on an existing T-N residual model. Inferred phenotypic groups also shared clinical characteristics with the training cohort. This indicates that T-N mismatch modeling is generalizable and therefore may have clinical utility and hold promise for personalized medicine. These T-N groups were also largely reproducible in the post-mortem cohort despite the limited regions of interest available and semiquantitative measurements of tau among other key differences with the *in vivo* data. Finally, similar, although not completely, overlapping phenotypes were also found using an alternative marker of neurodegeneration, ^18^F-fluorodeoxyglucose PET, in our prior work^22^.

Our study has some limitations. First, while we established T-N *mismatch* by modeling linear relationships between regional tau SUVR and thickness, the T-N relationship is likely to be, to some extent, non-linear even in pure AD cases. Non-linear approaches to measuring T-N *mismatch* using image to image translation may better model T-N relationships and capture additional spatial information. Second, our *ex vivo* validation on T-N *mismatch* utilized only six anatomical regions of interest due to specimen availability. We therefore missed information on the deviation of other regions when clustering. The semi-quantitative measures of tau histology further limit the sensitivity for assessment of deviations. Nonetheless, it is remarkable how similar the groups from this post-mortem dataset were to those in our *in vivo* analysis. An additional limitation to the histology is that PHF-1 is not specific to tau neurofibrillary tangles, so this may conflate other non-AD tauopathy contributions. Lastly, it is also worth noting that the cohorts used here were relatively modest in size and may not be generalizable to other cohorts, including those with greater numbers of co-morbidities and race/ethnicities that are not well represented in ADNI.

In summary, our findings demonstrated T-N *mismatch* depicts vulnerability and resilience likely attributable to specific non-AD pathologies or resilience factors. This approach may therefore provide important characterization of phenotypic heterogeneity in clinical populations, with implications for therapeutic trials and management.

## Methods

### Participants

#### ADNI Dataset

We included 343 participants from the Alzheimer’s Disease Neuroimaging Initiative (ADNI) dataset (http://adni.loni.usc.edu) who were classified with a diagnosis of mild cognitive impairment (MCI) or dementia. All participants had to have both a Tau PET scan and T1-weighted MRI scan. The closest MRI to tau PET scans were selected. The average time between tau scan and MRI scan was 14.4 (± 10) (SD) months. There were 184 amyloid positive (A+) and 159 amyloid negative (A-) patients included in this study. Most of the A+ patients were also in our prior study^21^. The summarized clinical characteristics of the cohort are reported in Table S4. In addition, we included 137 A-cognitively unimpaired adults from ADNI as controls in the voxel-wise thickness comparison analysis (Table S4).

#### AVID Dataset

We included 71 A+ symptomatic patients (36 female and 35 male) with a pair of Tau PET scan and T1-weighted MRI scans from the Avid Radiopharmaceuticals studies (A05) with inclusion criteria for age >=50 and MMSE > 10^45^. Participants provided written informed consent and both informed consent and the protocol were approved by the relevant Institutional Review Boards^45^. The Tau PET scan and T1-weighted MRI scan for each patient was obtained from the same visit. All participants were symptomatic, including 29 with dementia-level impairment and 42 with MCI. Average age was 73.6 ± 9.8, and average MMSE was 24.7 ± 4.3. The summarized clinical characteristics of this cohort was displayed in Table S5.

#### CNDR Dataset

We included 112 autopsies (age 72.0 ± 10 years at MRI scan and age 75.3 ± 11 years at death) from the University of Pennsylvania Center for Neurodegenerative Disease Research (CNDR). All individuals had an antemortem research-quality T1-weighted (T1w) MRI scan. For those with multiple scans, the closest to death was chosen. Semi-quantitative regional tau severity was determined by histology. The average time interval between MRI scan date and autopsy date was 46.0 (± 31) months. All procedures during life were performed with prior informed consent in accordance with Penn Institutional Review Board guidelines.

### Image acquisition and processing

#### Image acquisition

For both ADNI and AVID cohorts, we processed both T1-weighted MRI and tau PET (^18^F-flortaucipir) scans to obtain cortical thickness and tau SUVR for 104 bilateral gray matter regions of interest. The detailed image acquisition and processing methods have been previously described^21^. In brief, the T1w MRI scan of resolution 1.0×1.0×1.0 mm^3^ were acquired by ADNI, while PET images were of variable resolution, but reprocessed to a similar 0.8 cm full-width at half maximum resolution. FLAIR MRI was acquired in the same session as T1w MRI with variable spatial resolution as prescribed in the ADNI protocol. For the CNDR dataset, antemortem T1 structural MRI scan for all subjects were obtained with resolution ranging from 0.5×0.5×1mm^3^ to 1.25×1.25×1.20mm^3^. For AVID dataset, the MRI Scans have resolution 1.0×1.0×1.2 mm^3^.

#### T1-MRI processing

The image processing methods have been described in our prior work^21^. Briefly, the T1-weighted MRI was processed with the ANTs cortical thickness pipeline^46^ which includes steps for intensity inhomogeneity correction and tissue segmentation. The MRI scans were parcellated into cerebellar, cortical, and subcortical ROIs using a multi-atlas segmentation method^47^. The volumetric thickness map for each subject was estimated via DiReCT cortical thickness estimation method^48^ to generate volumetric thickness maps. ROI-based thickness was calculated by averaging the thickness maps across voxels within the gray matter ROIs. The same processing for T1-weighted MRI was applied to ADNI, AVID and CNDR cohorts.

#### PET processing

ADNI provides post-processed PET images that are generated by averaging co-registered individual frames. Post-processed PET images were registered to participants’ T1-weighted structural MRI using ANTs^49^. The following ANTs parameters was used. Metric: Mattes mutual information (weight=1, number of bins=32), Transformation model: Rigid (gradient step = 0.2), Smoothing levels = 4×2×0, Shrink factor = 4×2×1. MRI parcellated ROIs were transferred to PET space. Mean PET tracer uptake in cerebellar gray matter (18F-Flortaucipir) or cerebellar gray and white matter (18F-Florbetapir or 18F-Florbetaben) was used as a reference region to generate a standardized uptake value ratio (SUVR) map for each participation.

We additionally processed amyloid PET scans (18F-florbetaben or ^18^F-florbetapir tracer) for determining amyloid status. Amyloid status was determined by using a composite ROI measure of ^18^F-florbetaben or ^18^F-florbetapir tracer uptake^50^. As previously published^21^, we used an SUVR ≥ 1.11^41^ for ^18^F-Florbetapir and ≥ 1.08 for ^18^F-Florbetaben to define a positive amyloid scan (A+).

We processed 18F-Flortaucipir PET for AVID dataset. Attenuation-corrected image frames were first motion-corrected by MCFLIRT^51^ with 6 degree of freedom correction and averaged. The post-processed PET images were then processed following the same method as ADNI cohort to get regional SUVR.

#### White matter hyperintensities processing

The white matter hyperintensities (WMH) were segmented from FLAIR images using a deep learning-based method^52^ that was a top performer in a WMH segmentation challenge.

### Postmortem neuropathology measurement

All autopsies at the Penn CNDR were conducted with detailed procedures described elsewhere^53^, including routine examination of up to sixteen regions^53^ and uniform immunohistochemistry analyses. Briefly, the tissue was embedded in paraffin block, cut into 6 μm sections, and immunostained for a variety of proteins in specific regions. Antibody NAB228 was used to target amyloid deposits, PHF-1 to measure phosphorylated tau deposits and pS409/410 to detect phosphorylated TDP-43 deposits. The neuropathology burden of each region was then evaluated by pathologists by assigning a semi-quantitative score of none (0), rare (0.5), mild (1), moderate (2), or severe (3). Amyloid status was determined by histology amyloid score (C Score) with threshold at 2 based on prior work suggesting this threshold for the sensitivity of amyloid PET scans^35^.

### Modeling Tau (T) and Neurodegeneration (N) *mismatch* and clustering

#### A+ ADNI T-N mismatch clustering

The Tau (T) and neurodegeneration (N) relationship was modeled by robust linear regression between regional tau SUVR and cortical thickness, respectively. The bi-square weighting function was used to mitigate the effect of outliers. A natural log transformation was applied on tau SUVR as the independent variable to mitigate the effects of potentially skewed SUVR distribution. The regression residuals were discretized into a two-element binary vector based on whether they were more than 1.5 standard deviations away from the regression line. We used 1.5 standard deviation as the threshold “outliers”. In our prior work^21^, this threshold resulted in highly overlapping clusters compared to other thresholds or no threshold at all. These binarized vectors obtained from 104 bilateral regions of interest were entered into Ward’s D2 hierarchical clustering^23^ to generate data-driven grouping of subjects. The number of groups was determined by the elbow^24^ method which optimizes the within-group similarity and dendrogram structure. The three-dimensional regional mean residual map was visualized using MRIcroGL^54^.

#### A-ADNI thickness z-score clustering

The thickness of A-patients was standardized into z-score referenced to 137 cognitively normal individuals for all 104 cortical regions. Each regional z-score thickness was binarized based on 1.5 standard deviations. These binarized vectors obtained from 104 bilateral regions of interest were entered into Ward’s D2 hierarchical clustering^23^ to obtain N_Z_ groups in the same manner as above.

#### A+ AVID T-N mismatch testing

The regional T-N residuals of the AVID cohort were obtained from the regional T-N regression models built from the ADNI A+ cohort. Regional residuals were binarized based on the 1.5 standard deviation of residuals from the training set regression (ADNI). The Euclidean distance between each testing patient’s binarized vector and averaged binarized residual vector of T-N groups from ADNI cohort was obtained and compared. The group identity of each test patient was determined by finding the shortest Euclidean distance among all six A+ T-N groups obtained from ADNI.

#### *Ex vivo* T-N mismatch clustering

To validate T-N *mismatch* on *ex vivo* autopsies, we used cortical thickness measured from antemortem MRI and regional tau burden measured from histological staining for modeling T-N *mismatch*. Among all sixteen regions, only six cortical regions of interest without missing tau measurement were available from the postmortem samples: anterior cingulate gyrus, entorhinal cortex, angular gyrus plus middle occipital gyrus, middle frontal gyrus, superior temporal gyrus and amygdala. The T-N relationships were modeled following a procedure similar to the *in vivo* analysis, except that we additionally included the time between MRI scan date and autopsy date as covariate for modeling. The independent variable of tau burden was treated as a continuous variable rather than as a factor here since it resulted in lower Akaike information criterion (AIC)^55^, indicating better model fit, for all six regions. The same clustering procedure as used for the *in vivo* data was then performed on obtained residuals to partition subjects.

### ADNI comorbidities evaluation

Vascular risk factors data were obtained from the ADNI INITHEALTH table. Factors counted as vascular risk factors include hypertension, hyperlipidemia, type II diabetes, arrhythmia, cerebrovascular disease, endovascular management of head/neck vessels, coronary artery disease, coronary interventions, heart failure, structural heart defects/repair, peripheral artery disease, and smoking. The number of vascular risk factor for each patient was counted. Brain age was obtained by the machine-learning based Spatial Pattern of Atrophy for Recognition of Alzheimer’s Disease (SPARE) models^31^. The brain age gap was calculated by the difference between predicted brain age and the actual chronological age.

## Data Availability

All data produced in the present study may be available upon reasonable request to the authors

## Statistical analysis

Statistical analyses were performed in R (v4.5) or SPSS (v28). The between group comparison of continuous variables (e.g. regional tau SUVR) were analyzed by linear regression with covariates age and gender. Bonferroni correction was applied on all between-group comparisons. Comparison of ordinal or semi-quantitative variables (e.g., histology-measured TDP-43 severity levels) was conducted using Kruskal-Wallis tests^56^ corrected by multiple comparison or using Mann-Whitney test^57^ if only comparing between two groups. The voxel-wise thickness comparison was analyzed by using the threshold-free cluster enhancement method^58^ with age and gender as covariates. The global measures of and Clinical Dementia Rating Sum of boxes (CDRSB)^28^ were used to evaluate longitudinal cognitive changes. Longitudinal trajectories of cognitive scores were assessed with linear mixed-effects models^59^ using cognitive scores as the dependent variable. Fixed effects include time, group, and time*group interaction as predictors, and covariates (age, gender and years of education). A random intercept was included in the mixed-effects model to account for correlations among repeated measures of cognitive scores. The follow up time ranged from 1 to 4 years and maximum of 4 time points for each participant. Significant differences in rate of change between groups was determined by comparing the slope of time*group interaction. All statistical tests were two-sided.

## Acknowledgements

We acknowledge posthumously the significant contribution of John Q. Trojanowski to this work and the field of Alzheimer’s disease. The study was supported by the funding R01 AG056014 and RF1 AG069474. The CNDR brain bank was funded by NIH (P01AG066597, P30AG072979, U19AG062418). This study was also supported by MultiPark - A Strategic Research Area at Lund University. The AVID cohort in this work was sponsored by Avid Radiopharmaceuticals. Data collection and sharing for this project was funded by the Alzheimer’s Disease Neuroimaging Initiative (ADNI) (National Institutes of Health Grant U01 AG024904) and DOD ADNI (Department of Defense award number W81XWH-12-2-0012). ADNI is funded by the National Institute on Aging, the National Institute of Biomedical Imaging and Bioengineering, and through generous contributions from the following: AbbVie, Alzheimer’s Association; Alzheimer’s Drug Discovery Foundation; Araclon Biotech; BioClinica, Inc.; Biogen; Bristol-Myers Squibb Company; CereSpir, Inc.; Cogstate; Eisai Inc.; Elan Pharmaceuticals, Inc.; Eli Lilly and Company; EuroImmun; F.Hoffmann-La Roche Ltd and its affiliated company Genentech, Inc.; Fujirebio; GE Healthcare; IXICO Ltd.; Janssen Alzheimer Immunotherapy Research & Development, LLC.; Johnson & Johnson Pharmaceutical Research & Development LLC.; Lumosity; Lundbeck; Merck & Co., Inc.; Meso Scale Diagnostics, LLC.; NeuroRx Research; Neurotrack Technologies; Novartis Pharmaceuticals Corporation; Pfizer Inc.; Piramal Imaging; Servier; Takeda Pharmaceutical Company; and Transition Therapeutics. The Canadian Institutes of Health Research is providing funds to support ADNI clinical sites in Canada. Private sector contributions are facilitated by the Foundation for the National Institutes of Health (www.fnih.org). The grantee organization is the Northern California Institute for Research and Education, and the study is coordinated by the Alzheimer’s Therapeutic Research Institute at the University of Southern California. ADNI data are disseminated by the Laboratory for Neuro Imaging at the University of Southern California.

## Reference

1. Kapasi A, DeCarli C, Schneider JA. Impact of multiple pathologies on the threshold for clinically overt dementia. Acta Neuropathol (Berl). 2017;134(2):171–186. doi:10.1007/s00401-017-1717-7

2. Kapasi A, Schneider JA. Vascular contributions to cognitive impairment, clinical Alzheimer’s disease, and dementia in older persons. Biochim Biophys Acta. 2016;1862(5):878–886. doi:10.1016/j.bbadis.2015.12.023

3. Katsumata Y, Fardo DW, Kukull WA, Nelson PT. Dichotomous scoring of TDP-43 proteinopathy from specific brain regions in 27 academic research centers: associations with Alzheimer’s disease and cerebrovascular disease pathologies. Acta Neuropathol Commun. 2018;6(1):142. doi:10.1186/s40478-018-0641-y

4. Cummings JL. Cognitive and behavioral heterogeneity in Alzheimer’s disease: seeking the neurobiological basis. Neurobiol Aging. 2000;21(6):845–861. doi:10.1016/S0197-4580(00)00183-4

5. Lamar M, Boots EA, Arfanakis K, Barnes LL, Schneider JA. Common Brain Structural Alterations Associated with Cardiovascular Disease Risk Factors and Alzheimer’s Dementia: Future Directions and Implications. Neuropsychol Rev. 2020;30(4):546–557. doi:10.1007/s11065-020-09460-6

6. Robinson JL, Richardson H, Xie SX, et al. The development and convergence of co-pathologies in Alzheimer’s disease. Brain. 2021;144(3):953–962. doi:10.1093/brain/awaa438

7. Bartrés-Faz D, Arenaza-Urquijo E, Ewers M, et al. Theoretical frameworks and approaches used within the Reserve, Resilience and Protective Factors professional interest area of the Alzheimer’s Association International Society to Advance Alzheimer’s Research and Treatment. Alzheimers Dement Amst Neth. 2020;12(1):e12115. doi:10.1002/dad2.12115

8. Stern Y, Arenaza-Urquijo EM, Bartrés-Faz D, et al. Whitepaper: Defining and investigating cognitive reserve, brain reserve, and brain maintenance. Alzheimers Dement. 2020;16(9):1305–1311. doi:https://doi.org/10.1016/j.jalz.2018.07.219

9. Ossenkoppele R, Lyoo CH, Jester-Broms J, et al. Assessment of Demographic, Genetic, and Imaging Variables Associated With Brain Resilience and Cognitive Resilience to Pathological Tau in Patients With Alzheimer Disease. JAMA Neurol. 2020;77(5):632. doi:10.1001/jamaneurol.2019.5154

10. Lesuis SL, Hoeijmakers L, Korosi A, et al. Vulnerability and resilience to Alzheimer’s disease: early life conditions modulate neuropathology and determine cognitive reserve. Alzheimers Res Ther. 2018;10(1):95. doi:10.1186/s13195-018-0422-7

11. Whitwell JL, Josephs KA, Murray ME, et al. MRI correlates of neurofibrillary tangle pathology at autopsy: A voxel-based morphometry study. Neurology. 2008;71(10):743–749. doi:10.1212/01.wnl.0000324924.91351.7d

12. Wisse LEM, Ravikumar S, Ittyerah R, et al. Downstream effects of polypathology on neurodegeneration of medial temporal lobe subregions. Acta Neuropathol Commun. 2021;9(1):128. doi:10.1186/s40478-021-01225-3

13. Ravikumar S, Wisse LEM, Lim S, et al. Ex vivo MRI atlas of the human medial temporal lobe: characterizing neurodegeneration due to tau pathology. Acta Neuropathol Commun. 2021;9(1):173. doi:10.1186/s40478-021-01275-7

14. Joie RL, Visani AV, Baker SL, et al. Prospective longitudinal atrophy in Alzheimer’s disease correlates with the intensity and topography of baseline tau-PET. Sci Transl Med. 2020;12(524). doi:10.1126/scitranslmed.aau5732

15. Harrison TM, Joie RL, Maass A, et al. Longitudinal tau accumulation and atrophy in aging and alzheimer disease. Ann Neurol. 2019;85(2):229–240. doi:10.1002/ana.25406

16. Joie RL, Visani A, Bourakova V, et al. [P2–373]: AV1451-PET CORTICAL UPTAKE AND REGIONAL DISTRIBUTION PREDICT LONGITUDINAL ATROPHY IN ALZHEIMER’s DISEASE. Alzheimers Dement. 2017;13(7S_Part_15):P769–P769. doi:https://doi.org/10.1016/j.jalz.2017.06.1028

17. Das SR, Xie L, Wisse LEM, et al. Longitudinal and cross-sectional structural magnetic resonance imaging correlates of AV-1451 uptake. Neurobiol Aging. 2018;66:49–58. doi:10.1016/j.neurobiolaging.2018.01.024

18. Ossenkoppele R, Smith R, Ohlsson T, et al. Associations between tau, Aβ, and cortical thickness with cognition in Alzheimer disease. Neurology. 2019;92(6):e601–e612. doi:10.1212/WNL.0000000000006875

19. Jack CR, Bennett DA, Blennow K, et al. NIA-AA Research Framework: Toward a biological definition of Alzheimer’s disease. Alzheimers Dement J Alzheimers Assoc. 2018;14(4):535–562. doi:10.1016/j.jalz.2018.02.018

20. de Flores R, Wisse LEM, Das SR, et al. Contribution of mixed pathology to medial temporal lobe atrophy in Alzheimer’s disease. Alzheimers Dement J Alzheimers Assoc. 2020;16(6):843–852. doi:10.1002/alz.12079

21. Das SR, Lyu X, Duong MT, et al. Tau-Atrophy Variability Reveals Phenotypic Heterogeneity in Alzheimer’s Disease. Ann Neurol. 2021;90(5):751–762. doi:10.1002/ana.26233

22. Duong MT, Das SR, Lyu X, et al. Dissociation of tau pathology and neuronal hypometabolism within the ATN framework of Alzheimer’s disease. Nat Commun. 2022;13(1):1495. doi:10.1038/s41467-022-28941-1

23. Ward JH. Hierarchical Grouping to Optimize an Objective Function. Published online 1963. doi:10.1080/01621459.1963.10500845

24. Thorndike RL. Who belongs in the family. Psychometrika. Published online 1953:267–276.

25. Jack CR, Wiste HJ, Therneau TM, et al. Associations of Amyloid, Tau, and Neurodegeneration Biomarker Profiles With Rates of Memory Decline Among Individuals Without Dementia. JAMA. 2019;321(23):2316–2325. doi:10.1001/jama.2019.7437

26. Maass A, Landau S, Baker SL, et al. Comparison of multiple tau-PET measures as biomarkers in aging and Alzheimer’s disease. NeuroImage. 2017;157:448–463. doi:10.1016/j.neuroimage.2017.05.058

27. Upton J. Mini-Mental State Examination. In: Gellman MD, Turner JR, eds. Encyclopedia of Behavioral Medicine. Springer; 2013:1248–1249. doi:10.1007/978-1-4419-1005-9_473

28. O’Bryant SE, Waring SC, Cullum CM, et al. Staging Dementia Using Clinical Dementia Rating Scale Sum of Boxes Scores. Arch Neurol. 2008;65(8):1091–1095. doi:10.1001/archneur.65.8.1091

29. Cole JH, Marioni RE, Harris SE, Deary IJ. Brain age and other bodily “ages”: implications for neuropsychiatry. Mol Psychiatry. 2019;24(2):266–281. doi:10.1038/s41380-018-0098-1

30. Elliott ML, Belsky DW, Knodt AR, et al. Brain-age in midlife is associated with accelerated biological aging and cognitive decline in a longitudinal birth cohort. Mol Psychiatry. 2021;26(8):3829–3838. doi:10.1038/s41380-019-0626-7

31. Hwang G, Abdulkadir A, Erus G, et al. Disentangling Alzheimer’s disease neurodegeneration from typical brain ageing using machine learning. Brain Commun. 2022;4(3):fcac117. doi:10.1093/braincomms/fcac117

32. Moroni F, Ammirati E, Hainsworth AH, Camici PG. Association of White Matter Hyperintensities and Cardiovascular Disease. Circ Cardiovasc Imaging. 2020;13(8):e010460. doi:10.1161/CIRCIMAGING.120.010460

33. Nelson PT, Dickson DW, Trojanowski JQ, et al. Limbic-predominant age-related TDP-43 encephalopathy (LATE): consensus working group report. Brain. 2019;142(6):1503–1527. doi:10.1093/brain/awz099

34. Flores R de, Wisse LEM, Das SR, et al. Contribution of mixed pathology to medial temporal lobe atrophy in Alzheimer’s disease. Alzheimers Dement. 2020;16(6):843–852. doi:https://doi.org/10.1002/alz.12079

35. Mirra SS, Heyman A, McKeel D, et al. The Consortium to Establish a Registry for Alzheimer’s Disease (CERAD). Part II. Standardization of the neuropathologic assessment of Alzheimer’s disease. Neurology. 1991;41(4):479–486. doi:10.1212/wnl.41.4.479

36. Bakkour A, Morris JC, Wolk DA, Dickerson BC. The effects of aging and Alzheimer’s disease on cerebral cortical anatomy: Specificity and differential relationships with cognition. NeuroImage. 2013;76:332–344. doi:10.1016/j.neuroimage.2013.02.059

37. Latimer CS, Liachko NF. Tau and TDP-43 synergy: a novel therapeutic target for sporadic late-onset Alzheimer’s disease. GeroScience. Published online June 29, 2021. doi:10.1007/s11357-021-00407-0

38. Harper L, Bouwman F, Burton EJ, et al. Patterns of atrophy in pathologically confirmed dementias: a voxelwise analysis. J Neurol Neurosurg Psychiatry. 2017;88(11):908–916. doi:10.1136/jnnp-2016-314978

39. Huang W, Zhou Y, Tu L, et al. TDP-43: From Alzheimer’s Disease to Limbic-Predominant Age-Related TDP-43 Encephalopathy. Front Mol Neurosci. 2020;13. doi:10.3389/fnmol.2020.00026

40. Latimer CS, Burke BT, Liachko NF, et al. Resistance and resilience to Alzheimer’s disease pathology are associated with reduced cortical pTau and absence of limbic-predominant age-related TDP-43 encephalopathy in a community-based cohort. Acta Neuropathol Commun. 2019;7:9. doi:10.1186/s40478-019-0743-1

41. Duong MT, Nasrallah IM, Wolk DA, Chang CCY, Chang TY. Cholesterol, Atherosclerosis, and APOE in Vascular Contributions to Cognitive Impairment and Dementia (VCID): Potential Mechanisms and Therapy. Front Aging Neurosci. 2021;13:647990. doi:10.3389/fnagi.2021.647990

42. Neltner JH, Abner EL, Baker S, et al. Arteriolosclerosis that affects multiple brain regions is linked to hippocampal sclerosis of ageing. Brain. 2014;137(1):255–267. doi:10.1093/brain/awt318

43. Snyder HM, Corriveau RA, Craft S, et al. Vascular contributions to cognitive impairment and dementia including Alzheimer’s disease. Alzheimers Dement. 2015;11(6):710–717. doi:10.1016/j.jalz.2014.10.008

44. Lin Z, Sur S, Liu P, et al. Blood–Brain Barrier Breakdown in Relationship to Alzheimer and Vascular Disease. Ann Neurol. n/a(n/a). doi:10.1002/ana.26134

45. Devous MD, Fleisher AS, Pontecorvo MJ, et al. Relationships Between Cognition and Neuropathological Tau in Alzheimer’s Disease Assessed by 18F Flortaucipir PET. J Alzheimers Dis JAD. 2021;80(3):1091–1104. doi:10.3233/JAD-200808

46. Tustison NJ, Cook PA, Klein A, et al. Large-scale evaluation of ANTs and FreeSurfer cortical thickness measurements. NeuroImage. 2014;99:166–179. doi:10.1016/j.neuroimage.2014.05.044

47. Wang H, Suh JW, Das S, Pluta J, Altinay M, Yushkevich P. Regression-Based Label Fusion for Multi-Atlas Segmentation. Conf Comput Vis Pattern Recognit Workshop IEEE Comput Soc Conf Comput Vis Pattern Recognit Workshop. Published online June 20, 2011:1113–1120. doi:10.1109/CVPR.2011.5995382

48. Das SR, Avants BB, Grossman M, Gee JC. Registration based cortical thickness measurement. NeuroImage. 2009;45(3):867–879. doi:10.1016/j.neuroimage.2008.12.016

49. Avants BB, Epstein CL, Grossman M, Gee JC. Symmetric diffeomorphic image registration with cross-correlation: evaluating automated labeling of elderly and neurodegenerative brain. Med Image Anal. 2008;12(1):26–41. doi:10.1016/j.media.2007.06.004

50. Landau SM, Breault C, Joshi AD, et al. Amyloid-β imaging with Pittsburgh compound B and florbetapir: comparing radiotracers and quantification methods. J Nucl Med Off Publ Soc Nucl Med. 2013;54(1):70–77. doi:10.2967/jnumed.112.109009

51. Jenkinson M, Bannister P, Brady M, Smith S. Improved optimization for the robust and accurate linear registration and motion correction of brain images. NeuroImage. 2002;17(2):825–841. doi:10.1016/s1053-8119(02)91132-8

52. Kuijf HJ, Biesbroek JM, De Bresser J, et al. Standardized Assessment of Automatic Segmentation of White Matter Hyperintensities and Results of the WMH Segmentation Challenge. IEEE Trans Med Imaging. 2019;38(11):2556–2568. doi:10.1109/TMI.2019.2905770

53. Toledo JB, Van Deerlin VM, Lee EB, et al. A platform for discovery: The University of Pennsylvania Integrated Neurodegenerative Disease Biobank. Alzheimers Dement J Alzheimers Assoc. 2014;10(4):477-484.e1. doi:10.1016/j.jalz.2013.06.003

54. Rorden C, Brett M. Stereotaxic display of brain lesions. Behav Neurol. 2000;12(4):191–200. doi:10.1155/2000/421719

55. Bozdogan H. Model selection and Akaike’s Information Criterion (AIC): The general theory and its analytical extensions. Psychometrika. 1987;52(3):345–370. doi:10.1007/BF02294361

56. Kruskal WH, Wallis WA. Use of Ranks in One-Criterion Variance Analysis. J Am Stat Assoc. 1952;47(260):583–621. doi:10.2307/2280779

57. Mann HB, Whitney DR. On a Test of Whether one of Two Random Variables is Stochastically Larger than the Other. Ann Math Stat. 1947;18(1):50–60. doi:10.1214/aoms/1177730491

58. Smith SM, Nichols TE. Threshold-free cluster enhancement: addressing problems of smoothing, threshold dependence and localisation in cluster inference. NeuroImage. 2009;44(1):83–98. doi:10.1016/j.neuroimage.2008.03.061

59. nlme: Linear and nonlinear mixed effects models – ScienceOpen. Accessed February 17, 2022. https://www.scienceopen.com/document?vid=f71afca1-5770-4ec9-8abc-9f2da9326579

