## Supplemental Material for "Tau-Neurodegeneration *mismatch* reveals vulnerability and resilience to comorbidities in Alzheimer’s continuum"

* Co-corresponding authors.

### **Abstract**

Variability in the relationship of tau-based neurofibrillary tangles (T) and degree of neurodegeneration (N) in Alzheimer’s Disease (AD) is likely attributable to the non-specific nature of N, which is also modulated by such factors as other co-pathologies, age-related changes, and developmental differences. We studied this variability by partitioning patients within the Alzheimer’s continuum into data-driven groups based on their regional T-N dissociation, which reflects the residuals after the effect of tau pathology is “removed”. We found six groups displaying distinct spatial T-N *mismatch* and thickness patterns despite similar tau burden. Their T-N patterns resembled the neurodegeneration patterns of non-AD groups partitioned on the basis of z-scores of cortical thickness alone and were similarly associated with surrogates of non-AD factors. In an additional sample of individuals with antemortem imaging and autopsy, T-N mismatch was associated with TDP-43 co-pathology. Finally, T-N *mismatch* training was then applied to a separate cohort to determine the ability to classify individual patients within these groups. These findings suggest that T-N *mismatch* may provide a personalized approach for determining non-AD factors associated with resilience/vulnerability to Alzheimer’s disease.

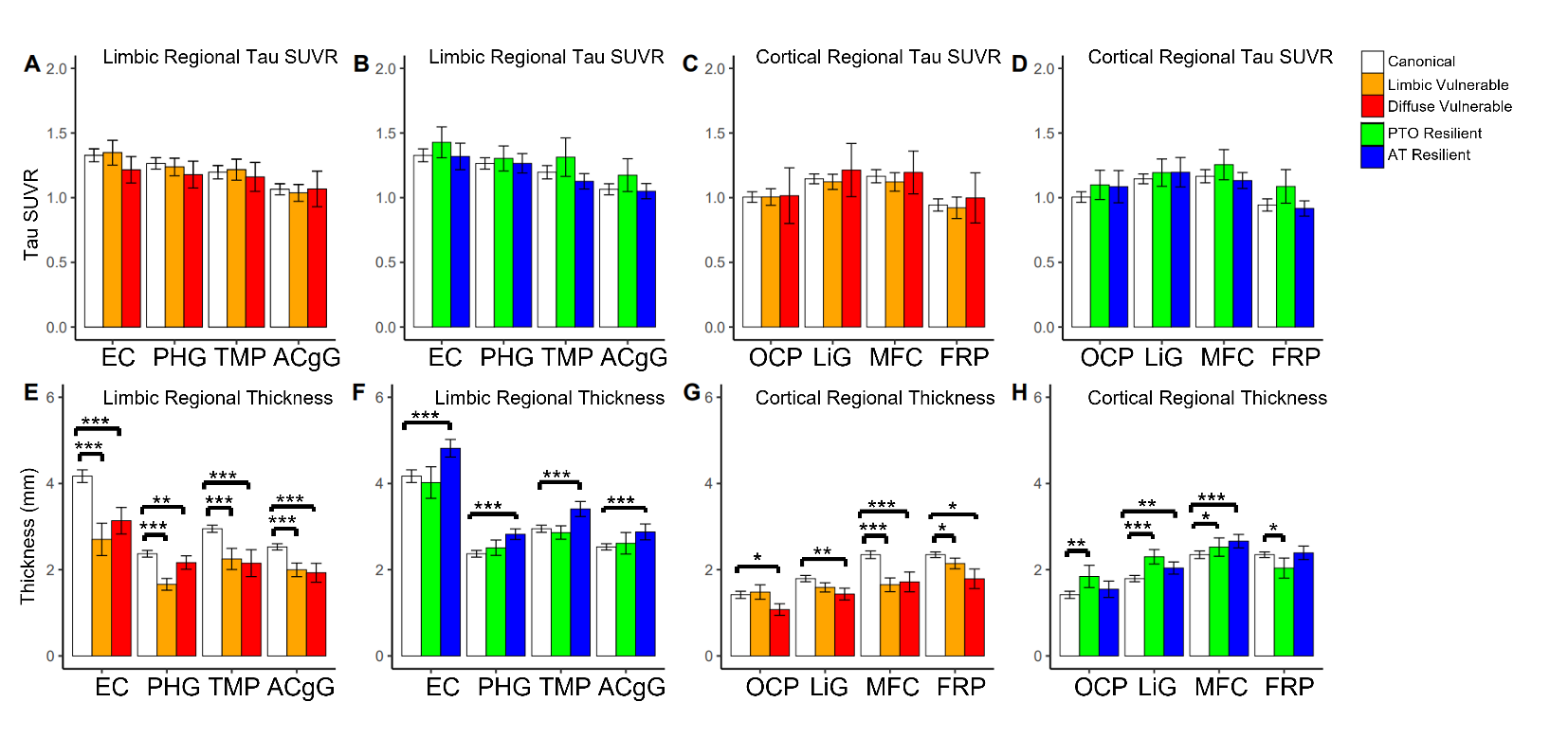
**Figure S1**. Top: Between-group comparisons of regional tau SUVR after covarying by age and sex for T-N groups (panels A, B, C, D). Bottom: Between-group comparisons of thickness across regions. The comparisons were controlling for age, associated regional tau SUVR and sex for T-N groups (panels E, F, G, H). Error bar represents ± 2 standard error. Abbreviations for regions of interest: EC = entorhinal cortex, PHG = parahippocampal gyrus, TMP = temporal pole, ACgG= anterior cingulate gyrus, OCP = occipital pole, LiG = lingual gyrus, MFC=medial frontal cortex, FRP=frontal pole. Significant levels after correction for multiple comparison are denoted as **P<0.05, **P<0.01, ***P<0.001*.

**Table S1.** Characteristics of distinct groups via N_Z_ clustering for 159 A- symptomatic patients on the basis of standardized regional thickness alone. Overall group effects are tested using the Kruskal-Wallis test for categorical variables (sex, MCI/Dementia) and linear regression for continuous variables (age, years of education, MMSE, CDRSB and inferior temporal (IT) tau SUVR). The baseline cognitive scores (Mini-Mental State Exam (MMSE)^20^, Clinical Dementia Rating Sum of boxes (CDRSB)^21^) and IT Tau SUVR was compared with age, sex and years of education as covariates. The mean (SD) is shown for age, years of education, MMSE, CDRSB and IT tau SUVR. Pairwise comparisons of these variables between N_Z_ groups were obtained. Only significant pairwise comparisons between T-N groups the Group1 (No Atrophy) were marked in the table. P-values were adjusted by multiple comparison correction (**P<0.05, **P<0.01, ***P<0.001)*.

| **Group**  **(n)** | **Description** | **Age** | **Sex**  **(F/M)** | **Diagnosis**  **(MCI/Dem)** | **Educ**  **(SD)** | **MMSE**  **(SD)** | **CDRSB**  **(SD)** | **IT Tau**  **SUVR**  **(SD)** |
| --- | --- | --- | --- | --- | --- | --- | --- | --- |
| Group1  (98) | No Atrophy | 73.8  (6.6) | 36/62 | 92/6 | 15.8  (3.1) | 28.3  (1.9) | 1.54  (1.6) | 1.15  (0.15) |
| Group2  (10) | Limbic Atrophy | 75.3  (9.0) | 2/8 | 6/4*** | 16.1  (2.6) | 24.9***  (3.6) | 4.45***  (3.8) | 1.15  (0.12) |
| Group3  (27) | Diffuse  Atrophy | 81.6***  (5.5) | 12/15 | 22/5 | 17.1  (2.5) | 26.7***  (3.6) | 2.40  (2.6) | 1.13  (0.091) |
| Group4  (24) | Posterior-Temporal Occipital  Increased Thickness | 67.0***  (5.5) | 12/12 | 24/0 | 16.0  (2.5) | 28.6  (1.4) | 1.09  (1.2) | 1.18  (0.21) |
| *Group Diff.* | -- | *P<0.001* | *P=0.352* | *P<0.001* | *P=0.246* | *P<0.001* | *P<0.001* | *P=0.589* |

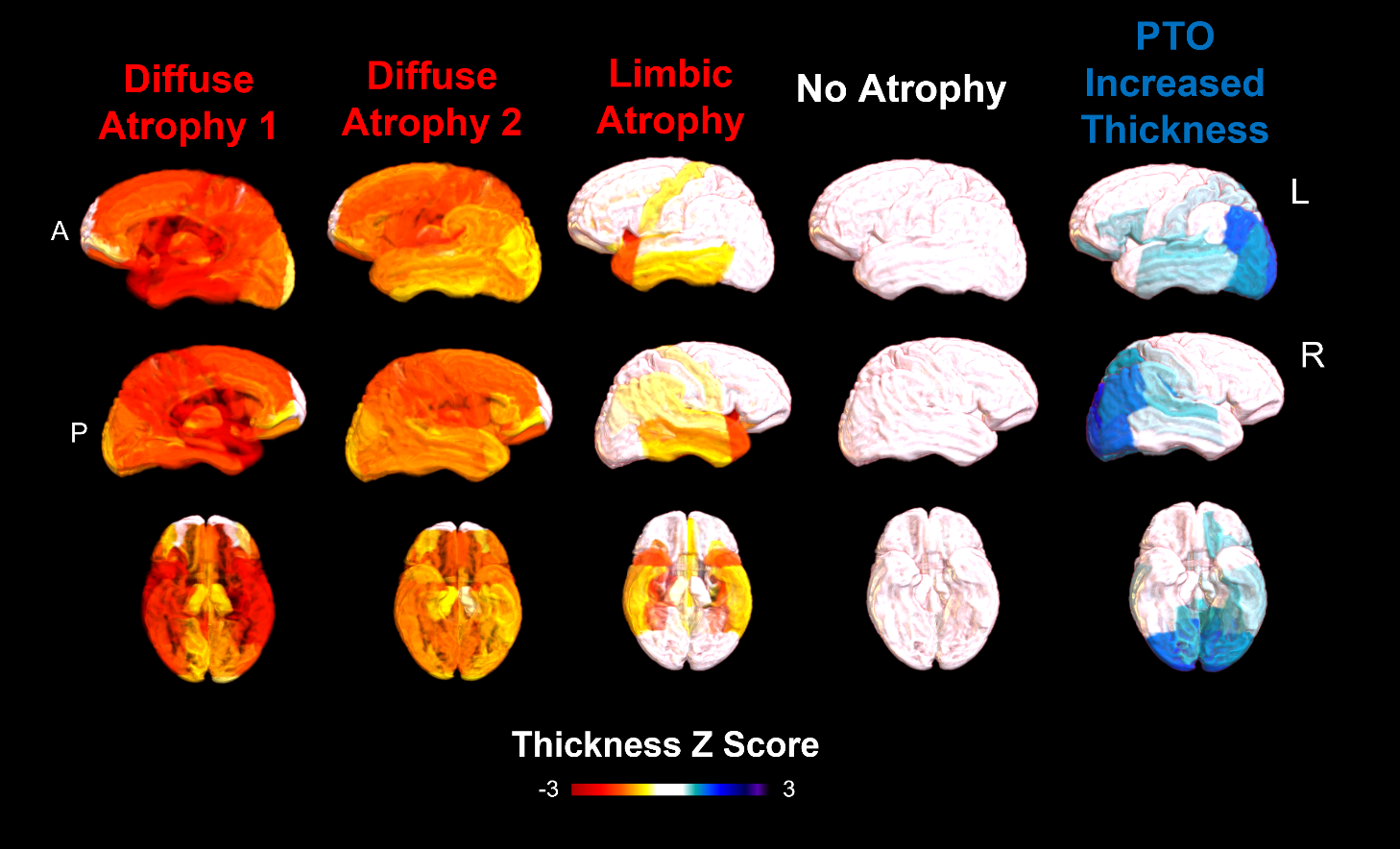
**Figure S2.** Regional thickness z-score for identified five N_Z_ groups among A- symptomatic patients from ADNI by clustering on standardized thickness using 137 normal individuals: no atrophy (close to 0 z-score), atrophy (negative z-score), increased thickness (positive z-score).

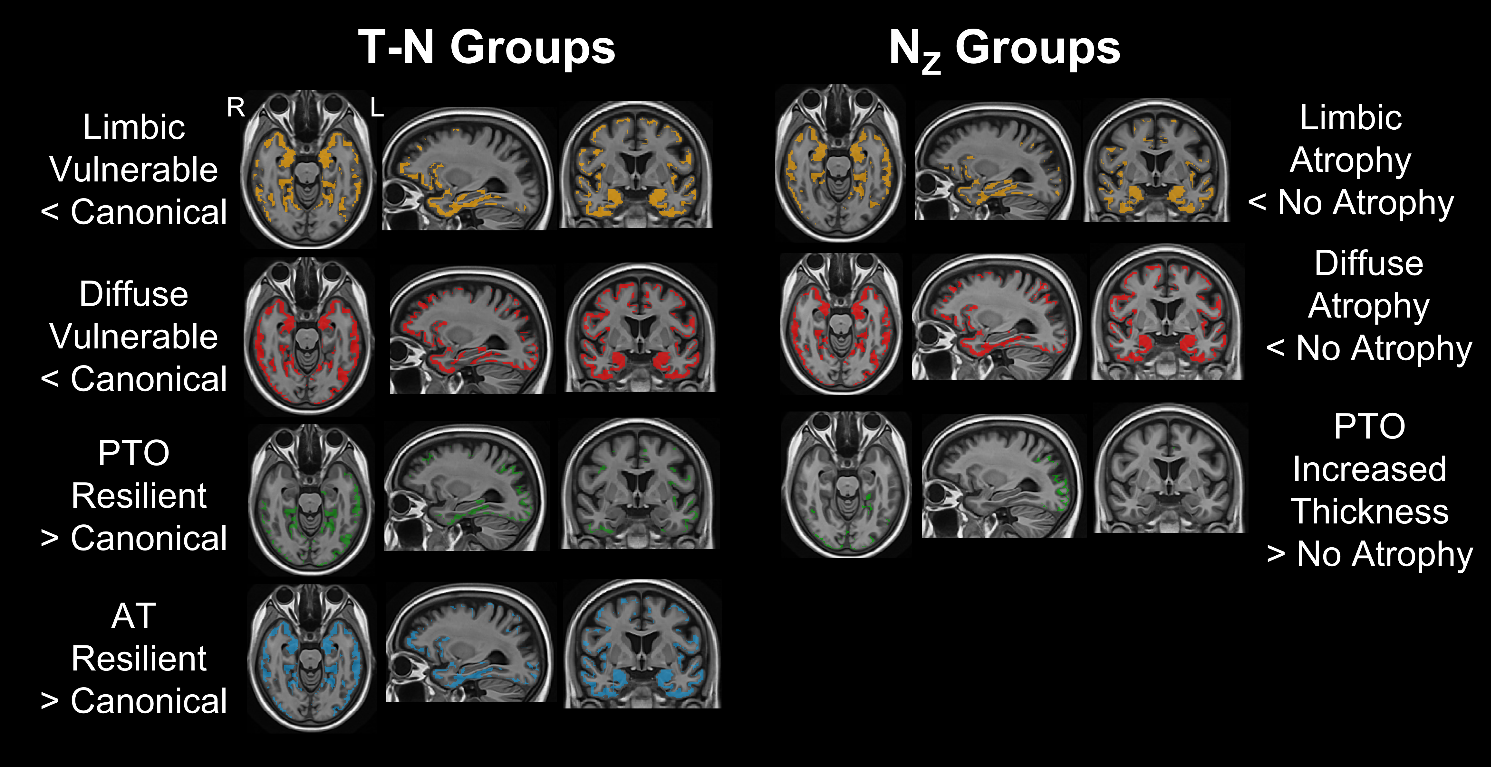
**Figure S3.** Voxel-wise significant differences of antemortem thickness between T-N vulnerable/resilient groups and the canonical group with *P_FWER_ < 0.05* for ADNI cohort (left). Voxel-wise significant differences of antemortem thickness between N_Z_ atrophy/increased thickness groups and the no-atrophy group with *P_FWER_ < 0.05* for ADNI cohort (right). The colored areas represent significant difference with the canonical for each group.

**Table S2.** Subjects Characteristics for T-N groups of 112 A+ symptomatic CNDR autopsies. Sex(F/M) was described in frequency. The mean (standard deviation) is shown for age at MRI scan and age at death. Significant pairwise comparison adjusted by multiple comparison with the canonical (Group1) was denoted as **P<0.05, **P<0.01, ***P<0.001*.

| **Group**  **(n)** | **Description** | **Sex**  **(F/M)** | **Age at MRI**  **(SD)** | **Age at Death**  **(SD)** |
| --- | --- | --- | --- | --- |
| Group1  (78) | Canonical | 52/26 | 71.8  (10) | 75.4  (11) |
| Group2  (11) | Limbic Vulnerable | 3/8 | 78.5  (10) | 81.1  (11) |
| Group3  (7) | Diffuse Vulnerable | 4/3 | 69.6  (11) | 71.1  (11) |
| Group4  (10) | Posterior-Temporal Occipital Resilient | 5/5 | 68.6  (9.0) | 71.5  (8.5) |
| Group5  (6) | Diffuse Resilient | 4/2 | 70.5  (7.5) | 73.8  (4.7) |
| *Group Diff.* | -- | *P=0.143* | *P=0.216* | *P=0.213* |

**Table S3.** Characteristics of inferred groups via T-N *mismatch* projection for 71 A+ symptomatic patients from AVID as testing cohort. Overall group effects are tested using the Kruskal-Wallis test for categorical variables (sex, MCI/Dementia) and linear regression for continuous variables (age, MMSE, ADAS-Cog11 and inferior temporal (IT) tau SUVR). The baseline cognitive scores (MMSE and ADAS-Cog11) and IT Tau SUVR was compared with age, sex and years of education as covariates. The mean (SD) is shown for age, MMSE, ADAS-Cog11 and IT tau SUVR. Pairwise comparisons of these variables between T-N groups were obtained. Only significant pairwise comparisons between T-N groups the Group1 (canonical) were marked in the table. P-values were adjusted by multiple comparison correction (**P<0.05, **P<0.01, ***P<0.001)*.

| **Cluster**  **(n)** | **Description** | **Age** | **Sex**  **(F/M)** | **Diagnosis**  **(MCI/Dem)** | **MMSE**  **(SD)** | **ADAS-Cog11**  **(SD)** | **ITG Tau**  **SUVR**  **(SD)** |
| --- | --- | --- | --- | --- | --- | --- | --- |
| Group1  (40) | Canonical | 72.7  (8.5) | 18/22 | 27/13 | 25.4  (3.9) | 13.1  (6.4) | 1.57  (0.42) |
| Group2  (12) | Limbic Vulnerable | 78.9  (9.6) | 7/5 | 3/9 | 21.2*  (4.7) | 24.1***  (9.5) | 1.69  (0.55) |
| Group3  (11) | Diffuse  Vulnerable | 78.1  (10.0) | 6/5 | 4/7 | 24.3  (4.5) | 16.7  (9.3) | 1.64  (0.41) |
| Group4  (5) | Posterior-Temporal Resilient | 62.2  (7.7) | 4/1 | 5/0 | 27.2  (2.0) | 16.6  (4.7) | 1.44  (0.40) |
| Group5  (2) | Anterior-Temporal  Resilient | 64.5  (13.4) | 1/1 | 2/0 | 26.0  (2.8) | 11.5  (6.4) | 1.99  (0.42) |
| Group6  (1) | Mixed | 82 | 0/1 | 0/1 | 23.0 | 15 | 1.25 |
| *Group Diff.* | -- | *P<0.01* | *P=0.639* | *P<0.01* | *P=0.019* | *P<0.001* | *P=0.483* |

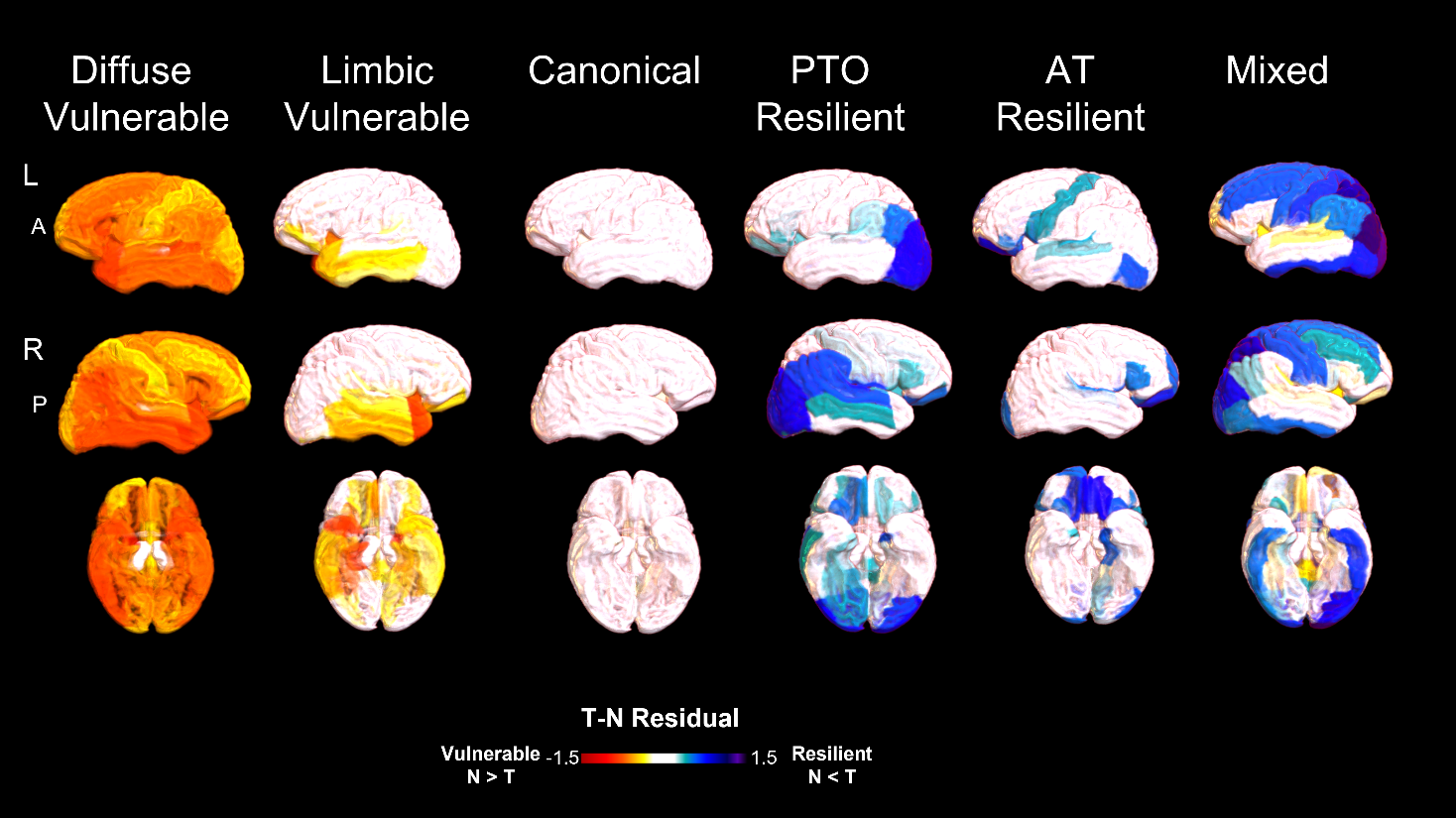
**Figure S4.** Average ROI-wise residual maps representing spatial T-N relationships for identified groups among A+ patients from AVID via testing on T-N *mismatch*: canonical (close to 0 residuals, N~T), vulnerable (negative residuals, N>T), resilient (positive residuals, N<T).

**Table S4.** Subjects Characteristics for 184 A+ symptomatic, 159 A- symptomatic and 137 cognitively normal ADNI subjects. The sex (F/M) and diagnosis status (MCI/Dementia/Normal) are described in frequency. The mean (standard deviation) is shown for age, years of education, MMSE and CDRSB.

| **Participants** | **Amyloid status** | **Age**  **(SD)** | **Sex**  **(F/M)** | **Educ**  **(SD)** | **MCI/**  **Dementia/**  **Normal** | **MMSE**  **(SD)** | **CDRSB**  **(SD)** |
| --- | --- | --- | --- | --- | --- | --- | --- |
| Symptomatic  patients | A+  (N=184) | 76.2  (8.0) | 103/81 | 15.8  (2.6) | 108/76/0 | 25.3  (3.9) | 3.00  (2.5) |
|  | A-  (N=159) | 74.2  (7.6) | 97/62 | 16.1  (2.9) | 144/15/0 | 27.8  (2.3) | 1.59  (1.5) |
| Normal  control | A-  (N=137) | 73.7  (6.7) | 75/62 | 16.8  (2.3) | 0/0/137 | 29.2  (1.1) | 0.0590  (0.25) |

**Table S5.** Subjects Characteristics for 71 A+ cognitively impaired AVID subjects. The sex (F/M) and diagnosis status (MCI/Dementia/Normal) are described in frequency. The mean (standard deviation) is shown for age, MMSE and ADAS-Cog11.

| **Participants**  **(n)** | **Age**  **(SD)** | **Sex**  **(F/M)** | **MMSE**  **(SD)** | **ADAS-Cog11**  **(SD)** |
| --- | --- | --- | --- | --- |
| MCI  (41) | 72.5  (9.4) | 18/23 | 27.2  (1.8) | 11.7  (4.7) |
| Dementia  (29) | 75.1  (10.4) | 18/11 | 21.1  (4.2) | 21.6  (8.9) |
